# Real-time prospective (shadow mode) validation of an AI-based clinical decision support system for predicting 3-month functional outcome in acute stroke: the VALIDATE study protocol

**DOI:** 10.64898/2026.04.26.26350937

**Authors:** Marta Rubiera, Martin Bendszus, Ronen R. Leker, Adam Hilbert, Ingo Werren, Luis M. Lopez-Ramos, Mercedes Ayesta, Thi Nguyet Que Nguyen, Sussane Bonekamp, Victoria Sala, Hamza Jubran, Claudia Meza, Fatma Shalabi, Yoel Schwartzmann, David Cano, Malte von Tottleben, John Kelleher, Dietmar Frey, the VALIDATE investigators

**Affiliations:** Hospital Universitari Vall d’Hebron, Stroke Unit, Barcelona, Spain; Vall d’Hebron Institut de Recerca, Stroke Research Group, Barcelona, Spain; University of Heidelberg, Department of Neuroradiology, Heidelberg, Germany; Hebrew University Hadassah Medical School, Department of Neurology, Yerushalayim, Israel; Charité Universitätsmedizin, Charité Lab for Artificial Intelligence in Medicine (CLAIM), Berlin, Germany; IBM iX GmbH, Digital Health Department, Berlin, Germany; Simula Metropolitan Center for Digital Engineering, Oslo, Norway; Rey Juan Carlos University – Fuenlabrada School of Engineering, Fuenlabrada, Spain; NORA Health, Sant Cugat, Spain; Technological University Dublin, Dublin, Ireland; Empirica Gesellschaft für Kommunikations- und Technologieforschung mbH, Digital Health, Bonn, Germany; Trinity College, ADAPT Research Centre, School of Computer Science and Statistics, Dublin, Ireland; Department of Neurosurgery, Charité – Universitätsmedizin Berlin, corporate member of Freie Universität Berlin and Humboldt-Universität zu Berlin, Germany

**Keywords:** Artificial intelligence, outcome prediction, acute stroke

## Abstract

**Introduction:** Despite the proven benefits of reperfusion therapies in acute ischemic stroke, treatment decisions in the hyperacute phase remain complex and are rarely supported by individualized outcome predictions. Artificial intelligence (AI)-based clinical decision support systems (CDSS) offer potential real-time prognostic estimates, but prospective evidence of their feasibility and performance in routine clinical workflows is limited. Our aim is to prospectively evaluate real-time feasibility, usability, and predictive performance of an AI-based CDSS (VALIDATE-CDSS) for individualized outcome prediction in acute stroke care.

**Methods and analysis:** Prospective, multicenter, observational study enrolling consecutive patients with acute ischemic stroke presenting to three tertiary stroke centers. Clinical management will follow standard practice at the discretion of treating physicians. In parallel, a dedicated researcher will collect patient data in real time and input them into the VALIDATE-CDSS using a mobile application, operating in shadow mode without influencing clinical decisions. The system will generate individualized predictions of 3-month functional outcome (modified Rankin Scale) for four treatment strategies (intravenous thrombolysis, endovascular thrombectomy, combined therapy, or no reperfusion) at three sequential time points: baseline clinical data, non-contrast CT, and CT angiography.

The primary outcome is the real-world feasibility and usability of the VALIDATE-CDSS in the hyperacute stroke workflow. Secondary outcomes include predictive performance, agreement between model-suggested and actual treatments, incremental value with increasing data availability, and assessment of potential bias across predefined subgroups. This study will provide prospective real-world evidence on the implementation and clinical potential of AI-based decision support for personalized treatment selection in acute ischemic stroke

**Ethics and dissemination:** Patient enrollment began after approval from the ethics committees of all participating centers. Results will be disseminated through peer-reviewed open-access journals and conference presentations. Following open science principles, anonymized data and metadata will be made publicly available in the Zenodo repository upon study completion.

**Trial registration:** ClinicalTrials.gov (NCT05622539).

**Strengths and limitations of this study:**

- First study to assess the feasibility of integrating an outcome-predictor CDSS into real-life hyperacute stroke workflows, addressing a critical gap between AI model development and clinical implementation
- Multicenter, prospective observational time-motion shadow-mode design, which minimizes interference with standard care while capturing real-world operational data
- Validation of a locked AI model developed from independent retrospective multicenter datasets across different populations, reducing the risk of overfitting to local case-mix
- Real-time data acquisition in the emergency department poses a significant operational challenge, with potential for missing or delayed inputs that may affect model performance in practice
- Risk of bias cannot be excluded, including spectrum bias from non-anticipated subgroups, temporal drift in clinical practice or patient populations, and centre-level variation in workflow and data quality

## Introduction

Artificial intelligence (AI)–based prognostic tools and clinical decision support systems (CDSS) can rapidly analyze large volumes of patient data to predict disease outcomes, supporting clinical decision-making. These tools enable more accurate and individualized treatment stratification, with the potential to improve patient outcomes and optimize healthcare resource use (1).

In acute ischemic stroke, the population-level benefits of intravenous thrombolysis (IVT) and mechanical endovascular thrombectomy (EVT) are well established (2). However, individual outcomes vary widely, and a considerable proportion of treated patients still experience poor recovery (3). Predicting outcomes in the hyperacute stroke setting is particularly challenging due to time constraints and the complex interaction of multiple prognostic factors (4). AI methods are well suited to this context, as they can identify complex, non-linear relationships across numerous variables and patient subgroups, and can be adapted for use across different levels of care (5).

VALIDATE, a Horizon Europe–funded project (No. 101057263) aims to test the feasibility of such AI integration into the acute stroke workflow. AI developers in the project together with Stroke Action For Europe patients’ association (SAFE) and clinical partners designed and created a machine learning based model to predict the modified Rankin Scale (mRS) (6) 3 months after stroke. The AI model has been fully developed and internally validated prior to this study using independent retrospective multicenter datasets from different populations, including retrospective data from the three comprehensive stroke centers (CSCs) participating in the present study (7). The model will remain locked during the study, with no further training or recalibration performed. For the present study, the model has been integrated into a CDSS combining structured data collection and real-time prediction.

We hypothesize that an AI-based CDSS providing individualized long-term outcome predictions can be feasibly integrated into real-world hyperacute stroke care workflows, and that it will demonstrate clinically meaningful predictive accuracy across the different hyperacute treatment options.

## Methods and analysis

### Study design

VALIDATE is a multicenter, prospective, phase 2 observational time-motion shadow-mode study conducted in three high-volume CSCs: two in Europe (Germany and Spain) and one in Israel. Consecutive acute ischemic stroke patients will be recruited over a 1.5-year period.

### Patient and Public Involvement Statement

The VALIDATE consortium includes the Stroke Action For Europe patients’ association (SAFE) as a full partner. SAFE has been involved since the earliest stages of project design, contributing particularly to the definition of outcomes that are meaningful to patients within a value-based healthcare framework. SAFE also plays a key role in dissemination, ensuring that results reach not only the clinical community but also healthcare institutions, patients, and their families.

### Patient population and study intervention

All patients admitted with acute ischemic stroke (<24 hours from symptom onset or stroke of unknown onset) will be screened for inclusion. Clinical management and treatment decisions will be performed according to local protocols and at the discretion of the treating stroke physicians.

After stroke code activation, a stroke-trained shadow researcher (SR) will accompany the treating team in the emergency department to enroll eligible patients, operating in a prospective time-motion “shadow mode” design. The SR will not display predictions or interact with treating clinicians, ensuring that the AI system does not influence clinical decision-making at any stage. Inclusion and exclusion criteria are detailed in Table 1.

**Table 1:** Inclusion and exclusion criteria of the prospective study.

| INCLUSION CRITERIA | EXCLUSION CRITERIA |
| --- | --- |
| - Age ≥ 18 years<br>- Acute ischemic stroke $<24$ hours from symptoms onset or unknown onset<br>- NIHSS $>1$<br>- Informed consent | - Known acute intracranial hemorrhage (ICH)<br>- Patients transferred from other facilities without repeated CT<br>- Serious, advanced, or terminal illness with anticipated life expectancy of less than 3 months<br>- Unlikely to be available for 90-day follow-up (e.g. no fixed home address, no telephone,...) |

Written informed consent will be obtained from the patient, a legal representative, a family member, or an independent physician when the patient is unable to consent. Re-consent will be sought during hospitalization if the patient later regains decision-making capacity.

For each enrolled patient, relevant clinical and imaging data will be entered in real time into a dedicated mobile application (VALIDATE app, see Figure 1).

**Figure 1:**
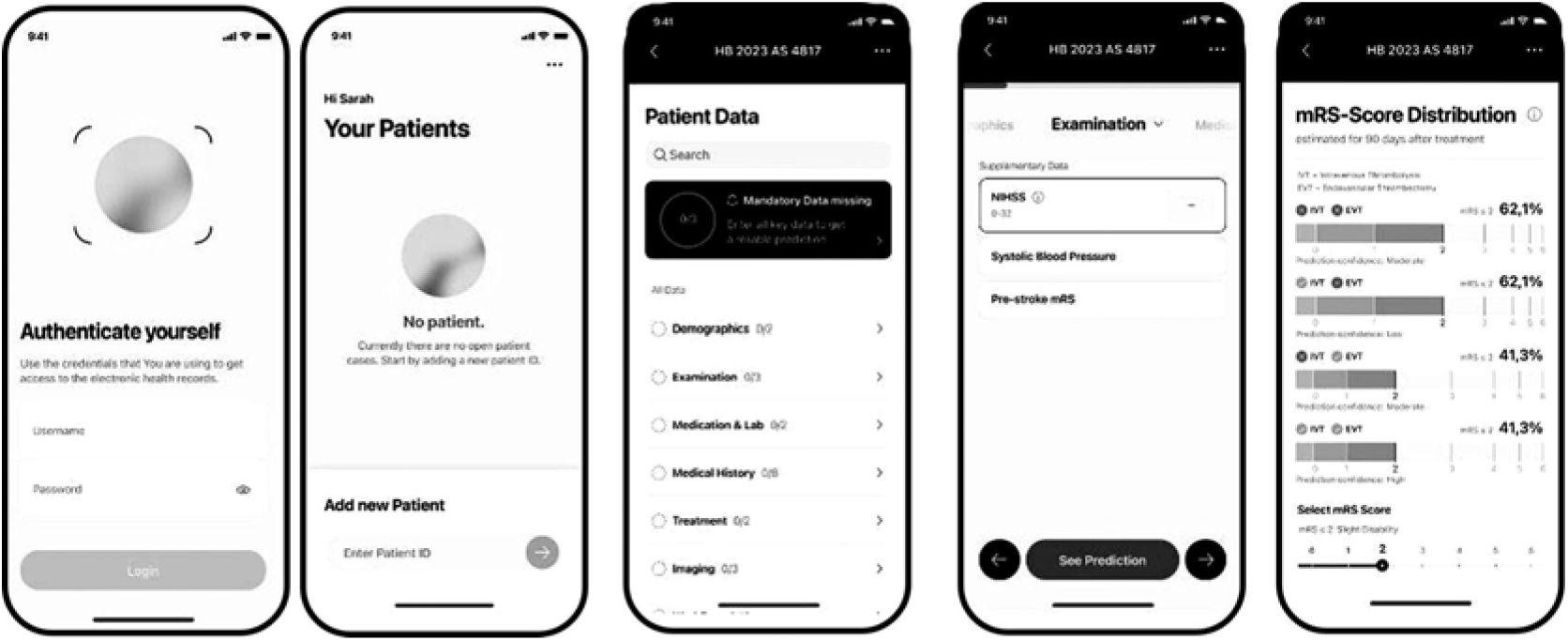
VALIDATE app: input data and prediction visualization.

Variables required for the AI prediction will be manually entered by the SR during the hyperacute phase without interfering with routine care. The VALIDATE-CDSS generates probabilistic predictions of the mRS at 3 months, even when some variables are missing. Predictions will be generated at three predefined time points along the hyperacute workflow (see Figure 2). After non-contrast computerized tomography (NCCT), patients with intracranial hemorrhage or non-ischemic pathology will be excluded from the study.

**Figure 2:**
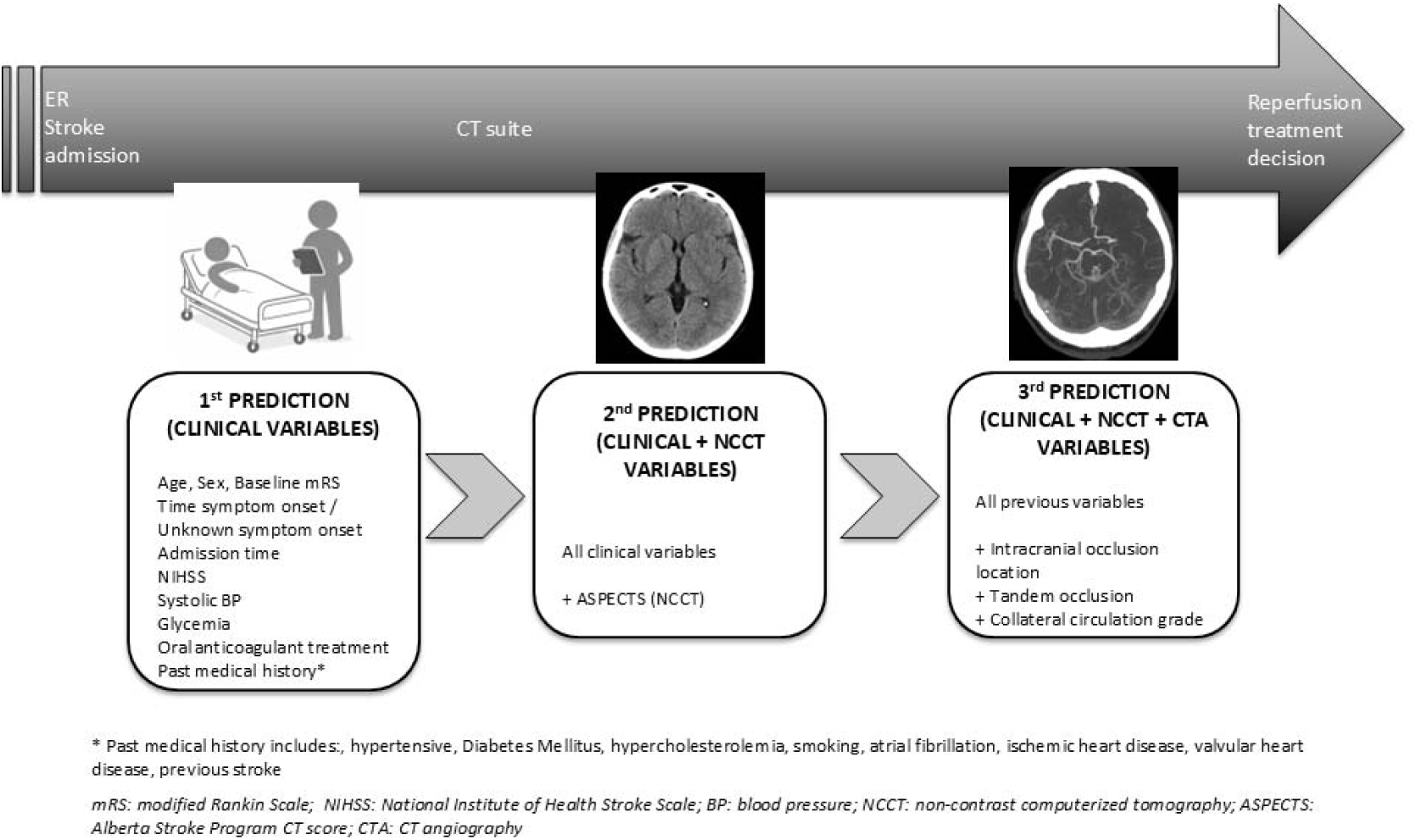
Workflow for sequential outcome prediction in the VALIDATE protocol.

Predictions will be manually triggered by the SR via the app. Data will be transmitted through a closed local wireless network to a dedicated workstation hosting the AI model, which will act as a backend server, and will enable triggering predictions and collecting data (Figure S1 from Online Supplemental). Prediction results will be returned and visualized in the app. This isolated local configuration complies with data protection requirements mandated by the ethics committees. In addition, the time-motion design will allow detailed characterization of workflow integration and operational impact of the AI system in the hyperacute setting.

The app displays outcome probabilities across the full mRS spectrum (scores 0–6) and as favorable/unfavorable outcomes using an adjustable threshold, stratified by treatment strategy: IVT, EVT, combined IVT+EVT, or no reperfusion treatment. Model confidence and visual representations of the contribution of different clinical variables to the predicted probability of treatment success (“data impact”) are also provided for individual predictions.

For each prediction, a structured report including predicted mRS probabilities, input variables, confidence metrics, and timestamps will be automatically generated and stored locally in the backend host workstation in CSV format.

Treating physicians will remain blind to all predictions. Patients will continue standard in-hospital care and follow-up. The actual mRS score at 3 months will be assessed by certified personnel, blinded to predictions, via telephone or in-person using standardized procedures. Additional outcomes, including patient-reported outcome measures (PROMs) will be collected from electronic medical records, telephone interviews, or through the NORA (8) digital platform, which enables patient education, communication, and survey-based PROM collection. A complete list of data collected for the study can be found in Table S.1 (Online Supplemental).

### Study endpoints

The primary objective is to assess the real-world feasibility and usability of real-time AI-based outcome prediction within the hyperacute stroke workflow.

Secondary objectives include evaluating the accuracy of AI predictions for 3-month functional outcome measured by the mRS across the full scale, dichotomized (mRS 0–2 vs. 3–6), and trichotomized (0–2, 3–4, 5–6) distributions. Additional analyses will assess concordance between predicted optimal treatment and actual treatment administered, correlation with PROMs, improvement in model performance with increasing data availability, and potential biases across predefined clinical and demographic subgroups.

### Data management and security

Given the observational design, no data safety monitoring board is required. A clinical research manager will oversee study conduct, with regular meetings and centralized monitoring through the NORA platform.

An electronic case report form (eCRF) integrated into NORA will store all study data. Acute-phase data collected by SR, outcomes, and key performance indicators will be consolidated into center-specific databases hosted on local hospital servers. After study completion, data will be anonymized locally and merged into a centralized dataset stored on secure servers at the Vall d’Hebron Institute of Research. All data handling complies with the European General Data Protection Regulation and relevant national regulations.

### Statistical analysis

Feasibility will be assessed by system penetration rate (target ≥70% during working hours), technical failures (<10% of technical failure during hyperacute phase), and door-to-prediction time. Usability will be evaluated using the System Usability Scale (target ≥68) and targeted surveys (see online supplemental Table S.2).

For the secondary outcomes, Cohen’s Kappa statistics will be used to measure the level of agreement between the treatment option with the better mRS predicted by the ML algorithms and real treatment received by the patient. Cohen’s Kappa will also be used to compare the outcome predicted by the VALIDATE-CDSS for the treatment received by the patient and his/her actual outcome.

Predictive performance will be primarily assessed using discrimination (AUROC), calibration, and agreement metrics, with additional classification metrics reported as secondary measures. Feature impact analysis will also be conducted. Following EU open science recommendations, anonymized data and metadata will be deposited in Zenodo after study completion. A complete statistical analysis plan can be found in the Supplemental Online Material.

### Sample size estimation

Sample size for this feasibility study was not determined by formal power calculation, as the primary aim is not to test a hypothesis but to estimate key feasibility parameters with sufficient precision to inform the design of a subsequent definitive trial. No formal hypothesis testing is planned for the primary endpoint.

A sample of 200 patients (approximately 67 per site over a recruitment period of 18 months) will allow estimation of the system penetration rate, the primary feasibility endpoint, with a 95% confidence interval of ±7 points around the expected target of 70% (based on a normal approximation for proportions). This precision is considered adequate to determine whether the pre-specified feasibility threshold has been met and to detect site-level variation in adoption. This approach is consistent with established guidance on the design and reporting of feasibility and pilot studies, which recommend that sample sizes be justified on the basis of estimation precision rather than statistical power.

### Ethics and Dissemination

The study is registered at ClinicalTrials.gov (NCT05622539), and patient enrollment started in July 2024, after approval from the Ethics Committees of the three clinical sites (Vall d’Hebron Institut de Recerca, PR(AG)432-2023; Universität Heidelberg Ethikkommission der Med. Fakultät, S-687/2023 and Helsinki Committee (Institutional Review Board) of Hadassah Medical Center-Ein Kerem, HMO-0529-22). This protocol was developed in accordance with the TRIPOD (9,10) guidelines for multivariable prediction models. All participating centers adhere to current European and international guidelines for acute stroke management (11). The protocol is publicly available in Zenodo (https://zenodo.org/records/17367485).

Results will be disseminated through peer-reviewed open-access journals and conference presentations. In line with open science principles, anonymized data and metadata will be made publicly available in the Zenodo repository upon study completion. The analytical code will be available after the future publication of the algorithms.

## Discussion

Predicting long-term outcomes for patients with acute ischemic stroke at emergency department admission remains highly challenging due to the interplay of multiple clinical factors, such as age, sex, pre-existing disability, stroke severity, time from symptom onset, and the variable effects of hyperacute treatments (12). Although randomized trials have demonstrated the efficacy of reperfusion therapies in selected populations, their strict inclusion criteria limit applicability in real-world settings (4). Consequently, clinical decision-making is often complex, especially for less experienced physicians or for patients who fall outside trial parameters. This complexity is intensified by the extreme time pressure of hyperacute care, leading to heterogeneous outcomes (13).

In this scenario, CDSS such as VALIDATE may assist clinicians by providing individualized outcome predictions under different treatment strategies. These tools can support real-time decisions and improve communication with patients and families by clarifying potential benefits and risks. Beyond acute care, CDSS may also enhance education, quality assurance, and continuous improvement of stroke services (14).

Previous predictive efforts have relied mainly on retrospective datasets and traditional statistical or AI-based models, typically offering dichotomized mRS predictions with accuracies between 0.70 and 0.80. Performance improves when incorporating treatment or early post-treatment data, but most models are developed in homogeneous cohorts and tested outside real-time workflows (15). In contrast, VALIDATE-CDSS was trained on large, multinational, multiracial datasets (7) and will be evaluated prospectively during hyperacute stroke care.

This study is the first to assess the feasibility of integrating a CDSS into real-life hyperacute workflows using an observational time-motion design, with a shadowing researcher accompanying the clinical team without interaction. This approach allows precise assessment of timing, integration, and usability while preserving the observational nature of the study by blinding physicians to predictions. Although shadowing methodologies are well established in behavioral sciences, their application to time-critical acute clinical workflows has been limited.

The target sample of 200 patients across three international centres over a recruitment period of 18 months reflects the expected volume of eligible acute stroke patients presenting during working hours at each participating site, based on historical admission rates. As this is a feasibility study, no formal power calculation was performed. The sample size is sufficient to assess system penetration, usability, and operational performance across diverse clinical settings, and to identify site-specific barriers to implementation that would inform the protocol of a future adequately powered trial (16).

A major challenge is real-time data acquisition, as some variables require bedside assessment and manual entry. VALIDATE-CDSS was therefore designed for rapid data input, local AI execution, and immediate result visualization without delaying treatment. The study will help identify potential workflow challenges introduced by AI-based CDSS.

## Summary

VALIDATE will evaluate the feasibility of a CDSS that provides individualized long-term outcome predictions for hyperacute ischemic stroke patients across different reperfusion strategies. If feasibility is demonstrated, these results will support the design of a randomized clinical trial comparing standard clinical decision-making with and without AI-based decision support.

## Author Contributions

Marta Rubiera: Conceptualization, Methodology, Supervision, Writing – Original Draft; Martin Bendszus: Conceptualization, Supervision, Writing – Review&Editing; Ronen R. Leker: Conceptualization, Supervision, Writing – Review&Editing; Adam Hilbert: Conceptualization, Funding Acquisition. Software, Validation, Writing – Review&Editing; Ingo Werren: Conceptualization, Software, Writing – Review&Editing; Luis M. Lopez-Ramos: Conceptualization, Software, Validation, Writing – Review&Editing; Mercedes Ayesta: Data Curation, Writing – Review&Editing; Thi Nguyet Que Nguyen: Software, Validation, Writing – Review&Editing; Sussane Bonekamp: Investigation, Writing – Review&Editing; Victoria Sala: Investigation, Writing – Review&Editing; Hamza Jubran: Investigation, Writing – Review&Editing; Claudia Meza: Investigation, Writing – Review&Editing; Fatma Shalabi: Investigation, Writing – Review&Editing; Yoel Schwartzmann: Investigation, Writing – Review&Editing; David Cano: Data Curation, Writing – Review&Editing; Malte von Tottleben: Project Administration, Writing – Review&Editing; John D. Kelleher: Conceptualization, Methodology, Writing – Review&Editing; Dietmar Frey: Conceptualization, Methodology, Supervision, Funding Acquisition.

All authors read and approved the final manuscript and agree to be accountable for all aspects of the work in ensuring that questions related to the accuracy or integrity of any part of the work are appropriately investigated and resolved.

## Acknowledgements

We thank all the VALIDATE consortium investigators and collaborators

