## Supplemental material for "Real-time prospective (shadow mode) validation of an AI-based clinical decision support system for predicting 3-month functional outcome in acute stroke: the VALIDATE study protocol"

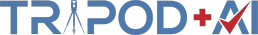
Version: 11-January-2024

| **_Section/Topic Item_ ^Development^ / evaluation**^1^ **Checklist item** | | | | **Reported**  **on page** |
| --- | --- | --- | --- | --- |
| **TITLE: Study protocol for the validation of a Trustworthy AI-based Clinical Decision Support System for Predicting Patient Outcome in Acute Stroke Treatment (VALIDATE)** | | | |  |
| *Title* | 1 | D;E | Identify the study as developing or evaluating the performance of a multivariable prediction model, the target population, and the outcome to be predicted | 1 |
| **ABSTRACT** | | | | |
| *Abstract* | 2 | D;E | See TRIPOD+AI for Abstracts checklist | 3-4 |
| **INTRODUCTION** | | | | |
| *Background* | 3a  3b  3c | D;E  D;E  D;E | Explain the healthcare context (including whether diagnostic or prognostic) and rationale for developing or evaluating the prediction model, including references to existing models  Describe the target population and the intended purpose of the prediction model in the context of the care pathway, including its intended users (e.g., healthcare professionals, patients, public)  Describe any known health inequalities between sociodemographic groups | 5  5  NA |
| *Objectives* | 4 | D;E | Specify the study objectives, including whether the study describes the development or validation of a prediction model (or both) | 5 |
| **METHODS** | | | | |
| *Data* | 5a  5b | D;E  D;E | Describe the sources of data separately for the development and evaluation datasets (e.g., randomised trial, cohort, routine care or registry data), the rationale for using these data, and representativeness of the data  Specify the dates of the collected participant data, including start and end of participant accrual; and, if applicable, end of follow-up | 5, 11  5, 10 |
| *Participants* | 6a  6b  6c | D;E  D;E  D;E | Specify key elements of the study setting (e.g., primary care, secondary care, general population) including the number and location of centres  Describe the eligibility criteria for study participants  Give details of any treatments received, and how they were handled during model development or evaluation, if relevant | 5, 6  Table 1  6 |
| *Data preparation* | 7 | D;E | Describe any data pre-processing and quality checking, including whether this was similar across relevant sociodemographic groups | NA |
| *Outcome* | 8a  8b  8c | D;E  D;E  D;E | Clearly define the outcome that is being predicted and the time horizon, including how and when assessed, the rationale for choosing this outcome, and whether the method of outcome assessment is consistent across sociodemographic groups  If outcome assessment requires subjective interpretation, describe the qualifications and demographic characteristics of the outcome assessors  Report any actions to blind assessment of the outcome to be predicted | 8  7  7 |
| *Predictors* | 9a  9b  9c | D  D;E  D;E | Describe the choice of initial predictors (e.g., literature, previous models, all available predictors) and any pre-selection of predictors before model building  Clearly define all predictors, including how and when they were measured (and any actions to blind assessment of predictors for the outcome and other predictors)  If predictor measurement requires subjective interpretation, describe the qualifications and demographic characteristics of the predictor assessors | 5 (Ref. 7)  Table S1, Figure 2  6 |
| *Sample size* | 10 | D;E | Explain how the study size was arrived at (separately for development and evaluation), and justify that the study size was sufficient to answer the research question. Include details of any sample size calculation | 8-9 |
| *Missing data* | 11 | D;E | Describe how missing data were handled. Provide reasons for omitting any data | NA |
| *Analytical methods* | 12a  12b  12c  12d  12e  12f  12g | D  D  D  D;E  D;E  E  E | Describe how the data were used (e.g., for development and evaluation of model performance) in the analysis, including whether the data were partitioned, considering any sample size requirements Depending on the type of model, describe how predictors were handled in the analyses (functional form, rescaling, transformation, or any standardisation).  Specify the type of model, rationale^2^, all model-building steps, including any hyperparameter tuning, and method for internal validation  Describe if and how any heterogeneity in estimates of model parameter values and model performance was handled and quantified across clusters (e.g., hospitals, countries). See TRIPOD-Cluster for additional considerations^3^  Specify all measures and plots used (and their rationale) to evaluate model performance (e.g., discrimination, calibration, clinical utility) and, if relevant, to compare multiple models Describe any model updating (e.g., recalibration) arising from the model evaluation, either overall or for particular sociodemographic groups or settings  For model evaluation, describe how the model predictions were calculated (e.g., formula, code, object, application programming interface) | Ref. 7  9 |
| *Class imbalance* | 13 | D;E | If class imbalance methods were used, state why and how this was done, and any subsequent methods to recalibrate the model or the model predictions | NA |
| *Fairness* | 14 | D;E | Describe any approaches that were used to address model fairness and their rationale | Ref. 7 |
| *Model output* | 15 | D | Specify the output of the prediction model (e.g., probabilities, classification). Provide details and rationale for any classification and how the thresholds were identified | 8, Ref.7 |

| *Training versus*  *evaluation* | 16 | D;E | Identify any differences between the development and evaluation data in healthcare setting, eligibility criteria, outcome, and predictors | 5, ref. 7, 5 |
| --- | --- | --- | --- | --- |
| *Ethical approval* | 17 | D;E | Name the institutional research board or ethics committee that approved the study and describe the participant-informed consent or the ethics committee waiver of informed consent | 10, 6 |
| **OPEN SCIENCE** | | | | |
| *Funding* | 18a | D;E | Give the source of funding and the role of the funders for the present study | 5 |
| *Conflicts of*  *interest* | 18b | D;E | Declare any conflicts of interest and financial disclosures for all authors | 12 |
| *Protocol* | 18c | D;E | Indicate where the study protocol can be accessed or state that a protocol was not prepared | 10 |
| *Registration* | 18d | D;E | Provide registration information for the study, including register name and registration number, or state that the study was not registered | 4 |
| *Data sharing* | 18e | D;E | Provide details of the availability of the study data | 9 |
| *Code sharing* | 18f | D;E | Provide details of the availability of the analytical code^4^ | ^10^ |
| **PATIENT & PUBLIC INVOLVEMENT** | | | | |
| *Patient & Public Involvement* | 19 | D;E | Provide details of any patient and public involvement during the design, conduct, reporting, interpretation, or dissemination of the study or state no involvement. | 5 |
| **RESULTS** | | | | |
| *Participants* | 20a  20b  20c | D;E  D;E  E | Describe the flow of participants through the study, including the number of participants with and without the outcome and, if applicable, a summary of the follow-up time. A diagram may be helpful. Report the characteristics overall and, where applicable, for each data source or setting, including the key dates, key predictors (including demographics), treatments received, sample size, number of outcome events, follow-up time, and amount of missing data. A table may be helpful. Report any differences across key demographic groups.  For model evaluation, show a comparison with the development data of the distribution of important predictors (demographics, predictors, and outcome). | 6-8 |
| *Model development* | 21 | D;E | Specify the number of participants and outcome events in each analysis (e.g., for model development, hyperparameter tuning, model evaluation) | Ref. 7 |
| *Model*  *specification* | 22 | D | Provide details of the full prediction model (e.g., formula, code, object, application programming interface) to allow predictions in new individuals and to enable third-party evaluation and implementation, including any restrictions to access or re-use (e.g., freely available, proprietary)^5^ | Ref. 7 |
| *Model*  *performance* | 23a  23b | D;E  D;E | Report model performance estimates with confidence intervals, including for any key subgroups (e.g., sociodemographic). Consider plots to aid presentation.  If examined, report results of any heterogeneity in model performance across clusters. See TRIPOD Cluster for additional details^3^. | Ref. 7 |
| *Model updating* | 24 | E | Report the results from any model updating, including the updated model and subsequent performance | Ref. 7 |
| **DISCUSSION** | | | | |
| *Interpretation* | 25 | D;E | Give an overall interpretation of the main results, including issues of fairness in the context of the objectives and previous studies | 10 |
| *Limitations* | 26 | D;E | Discuss any limitations of the study (such as a non-representative sample, sample size, overfitting, missing data) and their effects on any biases, statistical uncertainty, and generalizability | 11 |
| *Usability of the*  *model in the*  *context of current care* | 27a | D | Describe how poor quality or unavailable input data (e.g., predictor values) should be assessed and handled when implementing the prediction model | NA |
|  | 27b  27c | D  D;E | Specify whether users will be required to interact in the handling of the input data or use of the model, and what level of expertise is required of users  Discuss any next steps for future research, with a specific view to applicability and generalizability of the model | 12 |

From: Collins GS, Moons KGM, Dhiman P, et al. *BMJ* 2024;385:e078378. doi:10.1136/bmj-2023-078378

^1^ D=items relevant only to the development of a prediction model; E=items relating solely to the evaluation of a prediction model; D;E=items applicable to both the development and evaluation of a prediction model

^2^ Separately for all model building approaches.

^3^ TRIPOD-Cluster is a checklist of reporting recommendations for studies developing or validating models that explicitly account for clustering or explore heterogeneity in model performance (eg, at different hospitals or centres). Debray et al, BMJ 2023; 380: e071018 [DOI: 10.1136/bmj-2022-071018]

^4^ This relates to the analysis code, for example, any data cleaning, feature engineering, model building, evaluation

^5^ This relates to the code i Implement the model to get estimates of risk for a new individual.

**STATISTICAL ANALYSIS PLAN VALIDATE**

**Population**

The analysis will be performed in the per protocol population of acute ischemic stroke patients admitted as code strokes to the three comprehensive stroke centers included in the study. Patients not fulfilling the inclusion criteria or incurring any of the exclusion criteria (i.e no ischemic strokes) will be discarded during the emergency department evaluation and not considered for the study.

The algorithm is developed to suit real-life clinical scenarios. Therefore, handling missing data in terms of the variables used to obtain the prediction is integrated in its design. No missing data imputation procedures will be performed during the study. A testing of the incremental accuracy of the algorithm with an increasing number of variables is planned by computing predictions at 3 different time points for each patient: 1) exclusively with past medical history and clinical data recorded at admission of the stroke case, 2) adding non-contrast computerized tomography (CT) derived information and 3) adding CT angiography derived information.

No missing data imputation process is planned for the main functional outcome (modified Rankin scale at three months); data points where mRS90 is missing will be dropped (not taken into account for analysis). The calculated sample size includes an estimated 20% of loss to follow-up.

**Sample size estimation**

As the primary aim is feasibility rather than hypothesis testing, no formal a-priori sample size calculation was performed. Feasibility outcomes—including usability, integration into clinical workflows, data capture, and operational performance—are exploratory in nature and lack established parameters required for formal power calculations. However, an estimation based on expected predictive accuracy was conducted using Cohen’s Kappa for agreement between predicted and observed trichotomized mRS outcomes at 90 days.

Based on a retrospective cohort of 438 consecutive stroke patients from Hospital Vall d’Hebron (2020), mRS distribution was: 58.2% (0–2), 33.6% (3–4), and 8.1% (5–6). Assuming a minimum acceptable kappa of 0.6 and an expected kappa of 0.8, a sample size of 182 patients provides 95% power at a 0.05 significance level. Accounting for a 20% dropout rate, the target enrollment is 218 patients. Calculations were performed using the R package kappaSize.

**Primary and secondary outcomes**

As primary outcome, feasibility will be assessed by system penetration rate (target ≥70% during working hours), technical failures, and door-to-prediction time (target <30 minutes). Usability will be evaluated using the System Usability Scale (objective SUS >68) and targeted surveys (see online supplemental Table S.2), and a descriptive analysis will be performed, using mean +- standard deviation for normally distributed variables or median – interquartile range for those with non-normal distribution.

For the secondary outcomes, Cohen’s Kappa statistics will be used to measure the level of agreement between the treatment option with the best predicted mRS by the ML algorithms and real treatment received by the patient. Cohen’s Kappa will also be used to compare the outcome predicted by the VALIDATE-CDSS for the treatment received by the patient and the actual outcome.

Predictive performance will be assessed using weighted Cohen’s Kappa (with linear weights) to evaluate the level of agreement between the prediction by the ML-algorithms and the real outcome of the patients measured by the mRS at 3 months, considering the trichotomized distribution of mRS (0-2, 3-4, 5-6) and the full-scale (0-6). For the dichotomized outcome (0–2 vs. 3–6), standard (unweighted) Cohen’s Kappa will be used. A kappa ≥0.6 will be considered indicative of substantial agreement (Landis & Koch, 1977). All analysis of prediction performance will include evaluation of multiple metrics including precision, recall, f1-score, Receiver Operating Characteristic (ROC) as well as calibration plots (pre-/post adjustments with calibration methods such as Platt scaling) and bootstrapping for confidence intervals. Moreover, we will conduct bias analysis, including performance evaluation of patient sub-groups defined by clinical (e.g. treatment groups), demographic (e.g. ethnicity, sex) or robustness (outliers) criteria. Beyond predictions of the mRS score, assessment of differences in feature importance values (e.g. SHAP values) will be assessed. R and Python will be used to perform statistical analysis. An alpha level <0.05 (two-tailed) will be considered as statistically significant. All subgroup analyses are pre-specified and considered exploratory; no correction for multiple comparisons will be applied.

Following EU open science recommendations, anonymized data and metadata will be deposited in Zenodo after study completion and manuscript publication.

We will follow the Transparent Reporting of a multivariable prediction model for Individual Prognosis Or Diagnosis (TRIPOD+AI) guidelines to report our results (Collins G S et al. BMJ 2024; 385 :e078378 doi:10.1136/bmj-2023-078378).

### Table S1. VALIDATE variables by data source

| VALIDATE app  (predictors) | eCRF | NORA (or eCRF for non-NORA users) |
| --- | --- | --- |
| Age ^(1st)^ | Date of informed consent (ICF) signature | HADS – Depression score (90 days) |
| Biological sex^(1st)^ | Version of ICF | HADS – Anxiety score (90 days) |
| Time of symptom onset^(1st)^ | Gender | PROMIS10 Physical (7 days) |
| Admission time^(1st)^ | Ethnicity | PROMIS10 Mental (7 days) |
| NIHSS (baseline) ^(1st)^ | Thrombectomy (yes/no) | PROMIS10 Physical (90 days) |
| Systolic blood pressure^(1st)^ | Time of arterial puncture (EVT) | PROMIS10 Mental (90 days) |
| Pre-stroke mRS^(1st)^ | Intracranial occlusion location (DSA) | EQ-5D VAS (7 days) |
| Oral anticoagulants^(1st)^ | Baseline TICI | EQ-5D VAS (90 days) |
| Glycemia^(1st)^ | TICI after first pass |  |
| Hypertension^(1st)^ | Final TICI |  |
| Hypercholesterolemia^(1st)^ | Time of recanalization / end of procedure |  |
| Ischemic heart disease^(1st)^ | Thrombectomy complications |  |
| Smoking^(1st)^ | Hemorrhagic transformation (ICH) |  |
| Valvular heart disease^(1st)^ | Symptomatic ICH |  |
| Previous stroke^(1st)^ | NIHSS 24 h |  |
| Atrial fibrillation^(1st)^ | In-hospital complications |  |
| Diabetes mellitus^(1st)^ | NIHSS at 5 days or discharge |  |
| Imaging time^(2nd)^ | mRS at 5 days or discharge |  |
| ASPECTS^(2nd)^ | In-hospital mortality |  |
| Intracranial occlusion location ^(3rd)^ | mRS at 90 days |  |
| Collateral score ^(3rd)^ |  |  |
| Tandem occlusion^(3rd)^ |  |  |

^(1st):^ first prediction, past medical history and clinical data

^(2nd):^ second prediction, adding data from the non-contrast computerized tomography (CT

^(3rd):^ third prediction, adding data from the CT angiography.

App accounts for mobile application; eCRF, electronic case report form; HADS, Hospital Anxiety and Depression scale; PROMIS, patient-reported outcomes measurement information system; NIHSS, National Institute of Health Stroke Scale; EVT, endovascular (reperfusion) treatment; mRS, modified Rankin scale; DSA, digital subtraction angiography; EQ-5D VAS, European Quality of Life questionnaire Visual Analog Scale; TICI, thrombolysis in cerebral infarction; ICH, intracranial hemorrhage; ASPECTS, Alberta Stroke Program Early CT score.

**Table S.2:** System Usability Scale (SUS) and VALIDATE targeted surveys

*SUS*

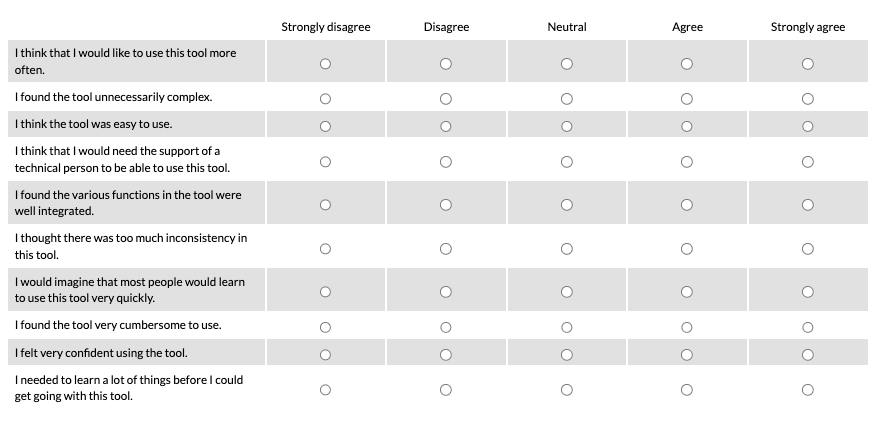

*VALIDATE targeted survey (research assistants after at least 5 initial patients included in the study)*

| Nr. | Question | Mandatory y/n | Question Type |
| --- | --- | --- | --- |
| 1 | May we contact you in the near future for a personal interview (duration 30 - 40 minutes)? The interview will take place remotely and will only serve to further evaluate your experience with the Validate CDSS). | Yes | Dichotomous (Yes/No) |
| 2 | Please give us your e-mail address where we can reach you. | No | Open-ended |
| 3 | How long have you been working in stroke treatment? | Yes | Single Choice |
| 4 | How would you rate your experience in stroke treatment? | Yes | Single Choice |
| 5 | In your opinion, how well can the Validate CDSS be integrated into the existing workflows of stroke care professionals? | Yes | Likert Scale (0-6) |
| 6 | What challenges do you think there are when integrating the Validate CDSS into everyday working life? | Yes | Multiple Choice + Open-ended |
| 7 | Compared to previous methods for decision making in stroke treatment - How helpful is the Validate CDSS for everyday work and decision making? | Yes | Likert Scale (0-6) |
| 8 | In which situations do you think the Validate CDSS is particularly useful? | No | Open-ended |
| 9 | In which situations do you think the Validate CDSS is a particular hindrance? | No | Open-ended |
| 10 | Can you imagine other use cases in which the Validate CDSS can be used in addition to support in the treatment of strokes? If yes, please describe the scenarios in a few words. | Yes | Open-ended |
| 11 | The following indications describe the intended use cases for the Validate CDSS  - I60-I69  - Acute ischemic stroke (I63)  - I60-I64  - ICH (intracranil hemorrhage)  - CVD (cardiovascular disease)  - TIA (transient ischemic atack)  Can you think of other indications for which the Validate CDSS can be used? If yes, please name the indication and explain your statement. | No | Open-Ended |
| 12 | The following contraindications describe all applications for which the Validate CDSS is not intended.  Contraindications:  - Hemorrhagic Stroke (I60-I62)  - Stroke mimics (any condition simulation an acute stroke)  - No evidence of clinical stroke  - Ischemic Stroke after 24 hours from known onset  - Acute stroke arriving to hospitals without reperfusion treatment as the algorithm is based on data from hospitals with reperfusion treatments available.  - No imaging evidence of stroke  - No evidence of clinical stroke  Can you think of any other contraindications for which the Validate CDSS cannot be used? | No | Open-Ended |
| 13 | Can you think of any social and mental factors (e.g. distractions, mental stress) and physical conditions (e.g. noise, temperature) that should be taken into account when using the service? | Yes | Open-ended |
| 14 | In your opinion, to what extent do the following user groups benefit from using the Validate CDSS in stroke treatment?   - Experts with extensive experience in stroke treatment - Expert with little experience in stroke treatment | Yes | Likert Scale (0-6) (2x) |
| 15 | Are there specific user groups for whom you think the Validate CDSS is particularly helpful? Please give reasons for your answer. | Yes | Dichotomous (Yes/No) + Open-ended |
| 16 | Are there certain user groups for whom you think the Validate CDSS is particularly unhelpful? Please give reasons for your answer. | Yes | Dichotomous (Yes/No) + Open-ended |
| 17 | How easy or complicated do you find the digital recording of the relevant patient data compared to the previous methods of (usual) documentation? If possible, give reasons for your answer. | Yes | Likert Scale (0-6) |
| 18 | How intuitive is the digital recording of the relevant data in the Validate CDSS for you? If possible, give reasons for your answer. | Yes | Likert Scale (0-6) |
| 19 | How clear and understandable is the presentation of the probabilities of success of the various treatment methods for you? Please give reasons for your choice if possible. | Yes | Likert Scale (0-6) |
| 20 | In this scenario, which of the possible treatment methods has the highest probability of success for a positive result 90 days after treatment? And which has the lowest? If possible, please give reasons for your choice. | Yes | Single Choice (2x) + Open-ended |
| 21 | In the Validate CDSS, you can see the probability of success of individual treatment methods and how the individual patient data influences this probability. Are you aware that this function exists and what its purpose is? | Yes | Dichotomous (Yes/No) |
| 22 | In your opinion, what is the reason that you are not aware that the module exists or what it is to be used for? | Yes | Open-ended |
| 23 | Please briefly summarize in your own words what the purpose of the function is and whether there is anything missing in the illustration. | Yes | Open-ended |
| 24 | How clearly does the Validate CDSS communicate to you that the probabilities of success calculated by the system are for decision support only, and should not and cannot replace your own clinical decision (or that of a colleague)? | Yes | Likert Scale (0-6) |
| 25 | Were there situations in which you had the feeling that the tool dictated or replaced a clinical decision? If yes, please give reasons for your answer. | Yes | Dichotomous (Yes/No) |
| 26 | How much do you trust the probabilities of success calculated by the Validate CDSS? If possible, please give reasons for your choice. | Yes | Likert Scale (0-6) |

*VALIDATE targeted survey (to be answered by stroke neurologist without previous contact with the VALIDATE app; screenshots and explanations of functionality will be provided)*

| Nr. | Question | Mandatory y/n | Question Type |
| --- | --- | --- | --- |
| 1 | How long have you been working in stroke treatment? | Yes | Single Choice |
| 2 | How would you rate your experience in stroke treatment? | Yes | Single Choice |
| 3 | What challenges are you currently experiencing in the collection and documentation of patient data in stroke treatment? What would you expect from a digital tool to support this process? | No | Open-ended |
| 4 | Do you understand what the purpose of “mandatory data” is? | Yes | Dichotomous (Yes/No) + Open-ended |
| 5 | Is there anything in the overview of probabilities that was unclear, irritating or surprising for you? | No | Dichotomous (Yes/No) + Open-ended |
| 6 | In your opinion, which mRS score should be pre-selected by default when the visualisation of results is initially loaded? Please give reasons for your choice, if possible. | No | Single Choice + Open-ended |
| 7 | Which treatment method would you choose based on the calculation of probabilities shown in the screenshot? | Yes | Open-ended |
| 8 | How helpful would you find a function that shows how individual patient data affects the probability of success of different treatment methods? | Yes | Likert Scale (0-6) |
| 9 | How much would you trust such a representation in a real clinical decision-making process? | Yes | Likert Scale (0-6) |
| 10 | To what extent do you consider the presentation of the factors influencing individual patient data on the probability of success to be suitable for helping less experienced colleagues (e.g. in further training) to understand clinical decisions in stroke treatment? | Yes | Likert Scale (0-6) |
| 11 | What additional information would you like to see in such a presentation to support your decision-making? | No | Open-ended |
| 12 | How clearly does the Validate CDSS communicate to you that the probabilities of success calculated by the system are for decision support only, and should not and cannot replace your own clinical decision (or that of a colleague)? | Yes | Likert Scale (0-6) |
| 13 | How much do you trust the probabilities of success calculated by the Validate CDSS? If possible, please give reasons for your choice. | Yes | Likert Scale (0-6) + Open-ended |
| 14 | In your opinion, how well can the Validate CDSS be integrated into the existing workflows of stroke care professionals? | Yes | Likert Scale (0-6) |
| 15 | What challenges do you think there are when integrating the Validate CDSS into everyday working life? | Yes | Multiple Choice + Open-ended |
| 16 | Compared to previous methods for decision making in stroke treatment - How helpful is the Validate CDSS for everyday work and decision making? | Yes | Likert Scale (0-6) |
| 17 | In which situations do you think the Validate CDSS is particularly useful? | No | Open-ended |
| 18 | In which situations do you think the Validate CDSS is a particular hindrance? | No | Open-ended |
| 19 | The following indications describe the intended use cases for the Validate CDSS  - ICD code: I60-I69  - Acute ischemic stroke (ICD-Code: I63)  - ICD code: I60-I64  - ICH (intracranial hemorrhage)  - CVD (cardiovascular disease)  - TIA (transient ischemic attack)  Can you think of other indications for which the Validate CDSS can be used? | No | Open-ended |
| 20 | The following contraindications describe all applications for which the Validate CDSS is not intended.  Contraindications:  - Hemorrhagic Stroke (I60-I62)  - Stroke mimics (any condition simulation an acute stroke)  - No evidence of clinical stroke  - Ischemic Stroke after 24 hours from known onset  - Acute stroke arriving to hospitals without reperfusion treatment as the algorithm is based on data from hospitals with reperfusion treatments available.  - No imaging evidence of stroke  - No evidence of clinical stroke  Can you think of any other contraindications for which the Validate CDSS cannot be used? | No | Open-ended |
| 21 | Can you think of any social and mental factors (e.g. distractions, mental stress) and physical conditions (e.g. noise, temperature) that should be taken into account when using the service? | Yes | Open-ended |
| 22 | Can you imagine other use cases in which the Validate CDSS can be used in addition to support in the treatment of strokes? If yes, please describe the scenarios in a few words. | Yes | Open-ended |
| 23 | In your opinion, to what extent do the following user groups benefit from using the Validate CDSS in stroke treatment?  - Experts with extensive experience in stroke treatment  - Expert with little experience in stroke treatment | Yes | Likert Scale (0-6) (2x) |
| 24 | Are there specific user groups for whom you think the Validate CDSS is particularly helpful? Please give reasons for your answer. | Yes | Dichotomous (Yes/No) + Open-ended |
| 25 | Are there certain user groups for whom you think the Validate CDSS is particularly unhelpful? Please give reasons for your answer. | Yes | Dichotomous (Yes/No) + Open-ended |
| 26 | Is there anything else you would like to share with us in connection with the use of an AI-based tool for decision support in stroke treatment? | No | Open-ended |
| 27 | If you are interested in expanding on your answers in a future interview, please provide your e-mail address. | No | Open-ended |

**Figure S1.** Data flow and management for the VALIDATE prospective study

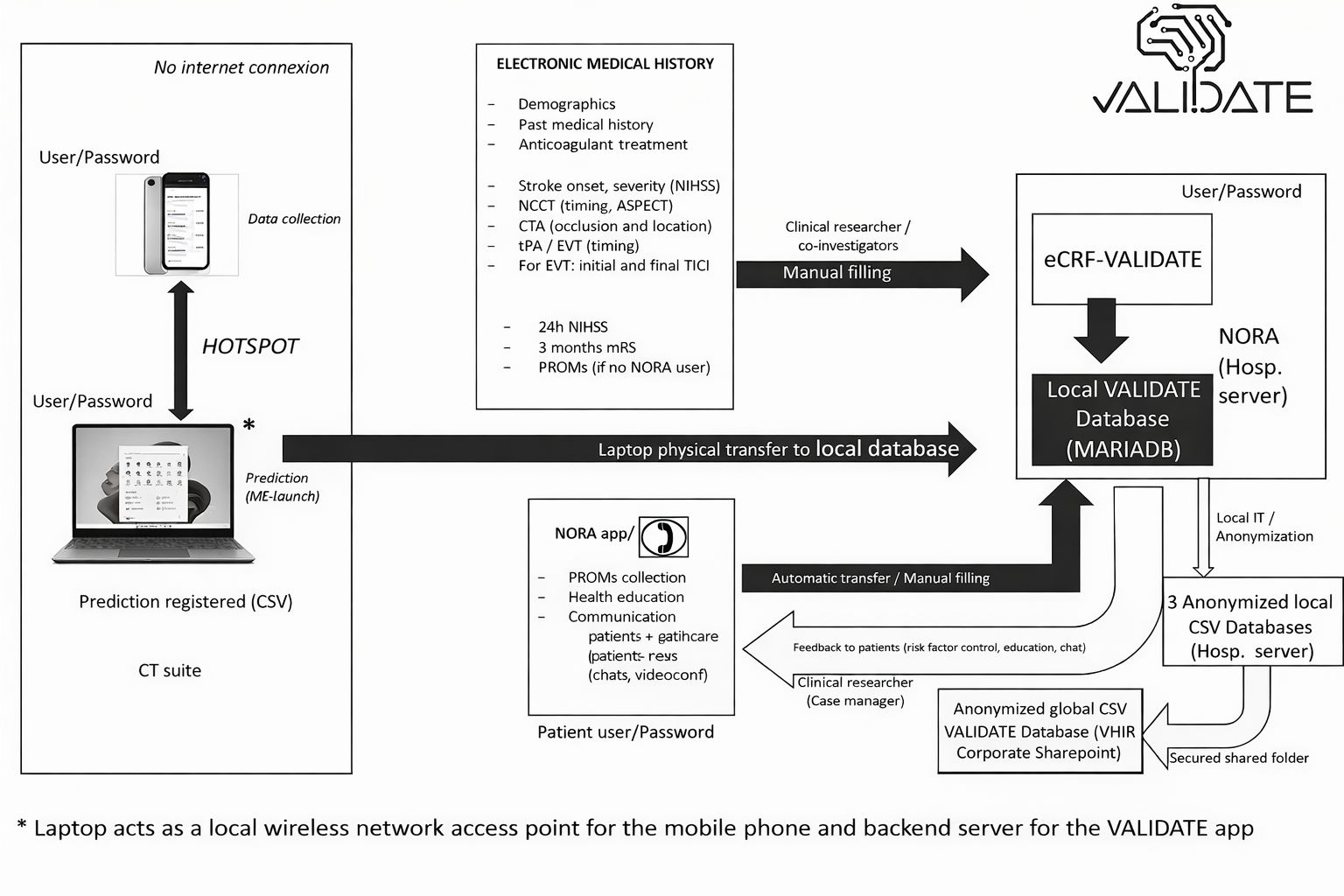
